# Correlation between the NRS-2002 score and PD-1/CTLA-4 levels in patients with CAP

**DOI:** 10.1101/2025.02.19.25322531

**Authors:** Chenguang Zhang, Mingqiang Zhang, Sheng Wu, Xiangdong Mu

## Abstract

**Objective:** To investigated the relationship between nutritional status and PD-1/CTLA-4 in CAP patients to determine whether the nutritional status is associated with the immunosuppression generated by T cells.

**Methods:** According the enrollment strategy, we enrolled 60 patients and collected their medical records.Take their blood samples and analyzed the distribution of PD-1 and CTLA-4 in different T cell subgroups.

**Results:** The level of PD-1 and CTLA-4 in the CD4^+^, CD8^+^ and Tregs were inclusive with the SCAP occurrence, mortality and PSI score.The malnutrition risk group suffered higher percentage of SAP, longer hospital-stay days and higher mortality when compared with the no-risk group.The higher the PD-1/CTLA-4 levels were, the higher the NRS-2002 score was.

**Conclusion:** A high NRS-2002 score may increase the length of hospital stay and the occurrence of SCAP. Higher PD-1 and CTLA-4 levels were associated with a higher PSI grade. Nutritional status influenced the occurrence of immunosuppression. Malnutrition status may increase the risk of immunosuppression, which is regulated by PD-1 and CTLA-4.

## 1. Introduction

Nutritional status have important influence on all populations, especially patients with comorbidities or critical diseases, elderly individuals, etc., and it is correlated with the prognosis and mortality of many diseases^1-3^. Research has shown that the prevalence of a high risk of malnutrition could reach 73% in elderly individuals in Asia^4^. In recognition of the increasing trends of hospitalization and mortality associated with community-acquired pneumonia (CAP) in modern times, especially after the Corona Virus Disease 2019 (COVID-19) pandemic, in-hospital mortality has reached more than 40%, and costs have increased sharply^5^.

Nutrient status affects infectious disease because it modulates inflammation and oxidative stress. Hypoproteinemia leads to low antibody production, including functional active immunoglobulins and gut-associated lymphoid tissue, and increases the risk of infection. When patients are invaded by a virus or bacteria, lymphocytes secrete cytokines and chemokines, triggering inflammation^6,7^. The nutrition risk screen-2002 (NRS-2002) is a classic, validated and fast method for evaluating nutritional status in the clinic. According to their NRS-2002 scores, patients can be separated into a malnutrition risk group (≥3) and a no-risk group (<3). Iddir M and Sümer A reported that nutritional status, as evaluated by the NRS-2002, affected the prognosis and severity of COVID-19 while the NRS 2002 score ≥3 group was at 5.6 times greater risk than the NRS 2002 score <3 group was (P = 0.0001, 95% CI = 66.636–1321.163) in terms of disease severity.^8^ However, few studies have investigated the relationship between CAP and nutritional status.

Several studies have noted that the immune response, which is mediated by T-cell subsets, including CD4^+^ T cells (CD4^+^), CD8^+^ T cells (CD8^+^), and regulatory T cells (Tregs), is correlated with the severity of infectious disease. PD-1 and CTLA-4, which are important T lymphocyte surface inhibitory molecules, are expressed on the surface of activated T lymphocytes^9-12^. Given that PD-1 and Cytotoxic T Lymphocyte antigen 4 (CTLA-4) expression is higher in patients who suffer from pneumonia or sepsis than in those in the control group, we propose that the duration of immunosuppression, which is regulated by CD4^+^, CD8^+^, and Tregs on the T-cell surface, plays an important role in the early stage of infection^10,12^.

Nutrients interact with anti-inflammatory cytokines via interactions with T cells. Malnutrition may induce immune deficiencies and result in a compromised immune response. However, there is limited evidence of a correlation between immunosuppression and nutritional status. Therefore, we explored the relationship between nutritional status and PD-1/CTLA-4 in CAP patients to determine whether the nutritional status is associated with the immunosuppression generated by T cells.

## 2. Materials and methods

### 2.1 Defnitions

The American Thoracic Society (ATS)/Infectious Diseases Society of America (IDSA) CAP guidelines address CAP as pneumonia acquired outside the hospital setting. Severe CAP (SCAP) was recognized according to the IDSA/ATS CAP severity criteria. The guidelines also strongly recommend that the pneumonia severity index (PSI) could be used as an effective clinical predictor for the prognosis of CAP. ^13-16^

In accordance with the European Society of Parenteral Enteral Nutrition guidelines, the NRS-2002 is a valuable assessment for screening nutritional status and dividing patients into a malnutrition risk group (score≥3 points) and a no-risk group (score<3 points).^17,18^

PD-1 and CTLA-4, which are expressed on the surface of CD4^+^, CD8^+^ and Treg cells, are considered indicators of immunosuppression that regulate T-cell function.

### 2.2 Study Design

#### 2.2.1 Enrollment Strategy

Enrollment criteria:1)patients agreed to take part in the research; 2)CAP was the main diagnose; 3)had complete clinical data;4)aged more than 18 years Exclusion criteria:1)agaed less than 18 years or preganace; 2)the clinical data uncomplete; 3)hospital acquired pneumonia (HAP); 4)CAP was the secondary doagnose; 5)COVID-19 infection pneumonia; 6) patients underwent the immnue-deficience disease or took the medication of immnue-depression treatment; 7)stop the research during the proceed.

#### 2.2.2 Data collection

This research was approved by the Ethics Committee of Beijing Tsinghua Changguang Hospital (BTCH) and was performed by the Emergency Department and Infectious Disease Center of BTCH.All patients agreed to participate in the research and draw blood samples for examination, and signed participant consent.

From Jan 2023 to Feb 2024,we screened 103 patients and enrolled 64 petients in the research according the enrollment and exclusion criteria. According the NRS-2002 scores,they were devided into malnutrition risk group (NRS-2002 scores≥3) and no-risk group (NRS 2002 scores<3).However during the proceed, 4 patients stop the research and transfered to other hospital or abandon the treatment. Finally 60 participants completed the research and divided into two groups according to their NRS-2002 scores.The malnutrition risk patients accounted 63.33% while the no-risk patients accounted 36.67% (38 vs. 22).The demographic information, comorbidities, laboratory parameters, PSI scores and NRS-2002 scores were collected through the hospital information system (HIS) of BTCH.(Figure 1)

**Figure 1.**
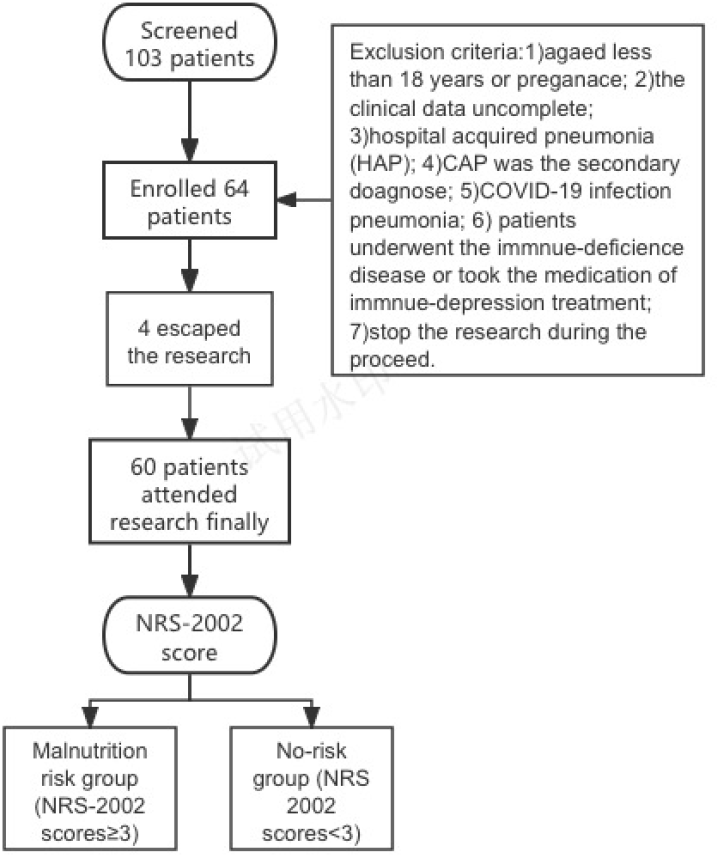
Enrollment strategy of the research

#### 2.2.3 Laboratory methods

Blood samples were collected within 24 hours from patients confirmed with CAP and stored in tubes containing dipotassium EDTA. All the samples were transferred at 4 °C within 2 hours, and the procedure was performed as follows:

1. Isolation of mononuclear cells in blood
2. Stimulate and block the secretion of lymphocytes
3. Direct immunofluorescence staining of cell surface
4. Intracellular direct immunofluorescence staining
5. FACS Canto II (BD Biosciences) was detected by streaming computer, and the results were analyzed by FCS EPress 4 (De Novo SoFtware).(Figure 2)

**Figure 2.**
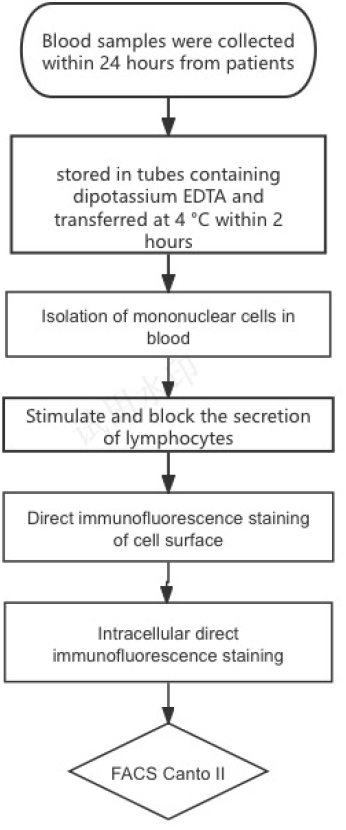
Laboratory methods

### 2.3 Outcomes

The occurance of SCAP and mortality was compared in different groups.The diagnose of SCAP was according the standards of IDSA/ATS CAP severity criteria. Mortality was considered to occur within 28 days after the enrollment of patients.

### 2.4 Statistical Analysis

Data analyzed by SPSS 30.0 and PRISM 10. The data with normal distribution were expressed as (X±s),T-test was used for comparison between them for comparison of the means of different groups. Non-normal distribution data were presented as M(P_25_,P_75_), the rank sum test was used for component comparison, and cross-tabulation was used for multi-group data analysis. Count data were expressed as a percentage of the number of cases. The correlation between data and patient survival was analyzed by survival analysis. The difference was statistically significant at P<0.05.

## 3. Results

### 3.1 Correlations between PD-1/CTLA-4 status and the severity of pneumonia

With respect to CD4^+^ PD-1, CD4^+^ CTLA-4, CD8^+^ PD-1, CD8^+^ CTLA-4, Tregs PD-1 and Tregs CTLA-4, the results of the logistic regression models concerning the association with SCAP occurrence and mortality were inconclusive (Table 1).

**Table 1.** Correlation between PD-1/CTLA-4 and SCAP and death.

|  | SCAP |  |  |  | Death |  |  |  |
| --- | --- | --- | --- | --- | --- | --- | --- | --- |
|  | Wals | P | OR | 95%CI | Wals | P | OR | 95%CI |
| NRS-2002 score | 3.98 | 0.04 | 4.18 | 1.02-17.04 | 2.54 | 1.11 | 0.12 | 0.01-1.63 |
| BMI(kg/m<sup>2</sup>)<sup>1</sup> | 0.72 | 0.40 | 1.91 | 0.79-1.78 | 0.07 | 0.80 | 1.07 | 0.65-1.77 |
| ALB(g/L) | 1.21 | 0.27 | 0.78 | 0.50-1.21 | 3.09 | 0.08 | 0.57 | 0.31-1.07 |
| CD4<sup>+</sup> PD1 | 0.54 | 0.46 | 1.15 | 0.80-1.65 | 0.60 | 0.44 | 1.19 | 0.79-1.83 |
| CD4<sup>+</sup>CTLA4 | 1.07 | 0.30 | 1.37 | 0.75-2.49 | 0.11 | 0.75 | 1.09 | 0.66-1.78 |
| CD8<sup>+</sup> PD1 | 1.32 | 0.25 | 1.15 | 0.91-1.45 | 0.83 | 0.36 | 0.89 | 0.70-1.13 |
| CD8<sup>+</sup>CTLA4 | 2.72 | 0.10 | 0.74 | 0.52-1.06 | 1.02 | 0.31 | 1.20 | 0.84-1.70 |
| Treg-PD1 | 0.02 | 0.89 | 1.02 | 0.75-1.39 | 2.66 | 0.10 | 1.28 | 0.95-1.71 |
| Treg-CTLA4 | 0.32 | 0.57 | 0.89 | 0.59-1.34 | 0.27 | 0.60 | 0.91 | 0.64-1.63 |

1.BMI=body mass index

Except those for PD-1 and CTLA-4 on CD4 (P1=0.01, P2<0.01), the correlations of CD4^+^ PD-1, CD4^+^ CTLA-4, CD8^+^ PD-1, CD8^+^ CTLA-4, Tregs PD-1 and Tregs CTLA-4 with PSI were not statistically significant (Table 2).

**Table 2.**
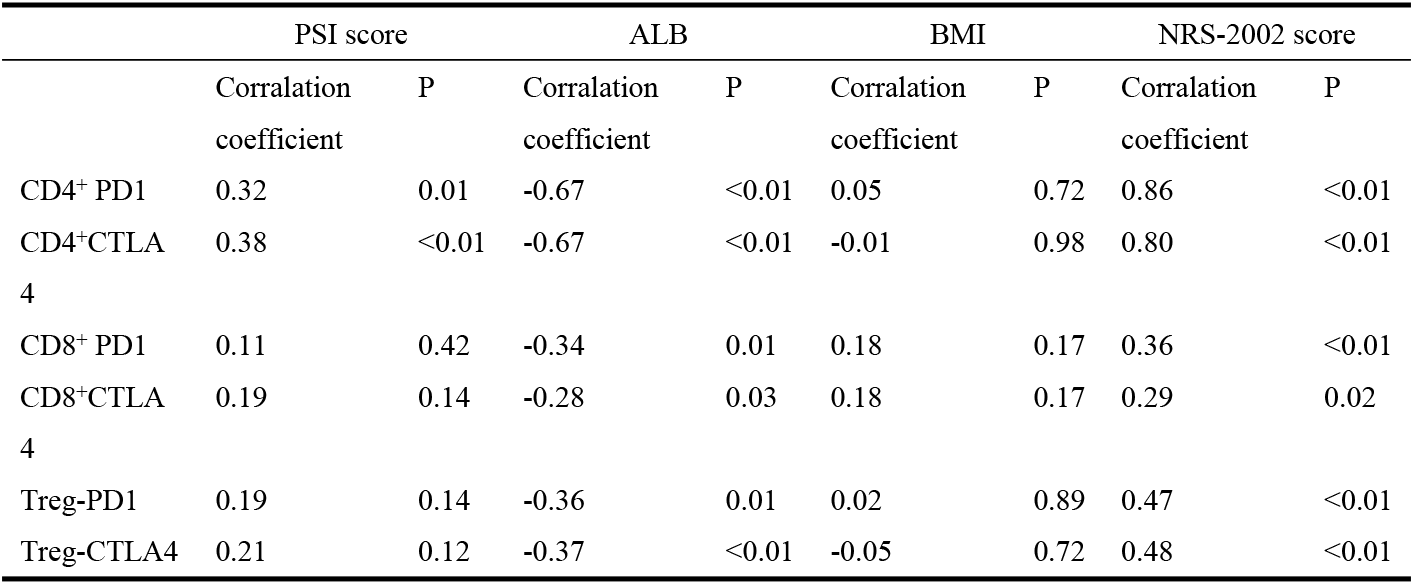
Correlation of PD-1/CTLA-4 and PSI, ALB, BMI and NRS-2002.

### 3.2 Influence of nutritional status on pneumonia

According to the inclusion criteria, 60 participants were enrolled and divided into a malnutrition risk group and a no-risk group (38 vs. 22). The NRS-2002 score in the malnutrition risk group was 4.02±1.21, whereas that in the no-risk group was 1.54±0.50. No significant differences between the two groups were confirmed for sex, age or comorbidities at baseline. Hospital mortality in the malnutrition risk group was greater than that in the no-risk group (23.68% vs. 4.54%). For the outcomes of hospital-free days and the occurrence of SAP, the no-risk group was prioritized over the malnutrition risk group (Table 3).

**Table 3.** Comparison between the different nutritional group.

|  | No-risk group | Malnutrition risk group | Value | P |
| --- | --- | --- | --- | --- |
| Number | 22 | 38 | - | - |
| Age(years old) | 63.40±19.19 | 69.57±19.18 | 1.20 | 0.23 |
| Male | 10(45.5) | 20(52.6) | 0.29 | 0.59 |
| Heart disease | 6(27.3) | 11(28.9) | 0.02 | 0.89 |
| Diabetes | 7(31.8) | 15(39.5) | 0.35 | 0.55 |
| CKD<sup>1</sup> | 2(0.9) | 8(21.1) | 1.43 | 0.23 |
| Mental disease | 4(18.1) | 10(26.3) | 0.51 | 0.47 |
| NRS-2002 scores | 1.54±0.50 | 4.02±1.21 | 9.07 | <0.01 |
| CRP(mg/L)<sup>2</sup> | 47.60(13.92,104.50) | 33.00(11.00,88.00) | 367.50 | 0.44 |
| PCT(ng/L)<sup>3</sup> | 2.80(1.22,4.32) | 8.80(3.20,18.92) | 662 | <0.01 |
| PSI score | 3.50(2.00,4.00) | 3.00(2.22,4.00) | 408.50 | 0.88 |
| Length of hospitalization(day) | 11(8.00,16.50) | 15.00(13.50,20.50) | 450 | <0.01 |
| SCAP | 1(0.5) | 13(34.2) | 10.6 | <0.01 |
| Death | 1(0.5) | 9(23.7) | 3.67 | 0.06 |

1.CKD=choicr kindey disease;2.CRP=C-reactive protein;3.PCT=Procalcitonin

For the NRS-2002 score, BMI and ALB, the OR results from the logistic regression models for SCAP and death, the NRS-2002 score was associated with the occurrence of SCAP. None of these factors were related to the risk of death (Table 1).

### 3.3 Distribution of PD-1/CTLA-4 in patients with different nutritional statuses

After CD4^+^ PD-1, CD4^+^ CTLA-4, CD8^+^ PD-1, CD8^+^ CTLA-4, Tregs PD-1 and Tregs CTLA-4 were analyzed in the two groups, the malnutrition risk group presented higher levels than those in the no-risk group. PD-1/CTLA-4 expression in the CD4^+^, CD8^+^ and Tregs subgroups was positively correlated with the NRS-2002 score. The higher the PD-1/CTLA-4 levels were, the higher the NRS-2002 score was. Similar results were obtained for the ALB concentration and BMI, which are essential for evaluating nutritional status (Figure 3).

**Figure 3.**
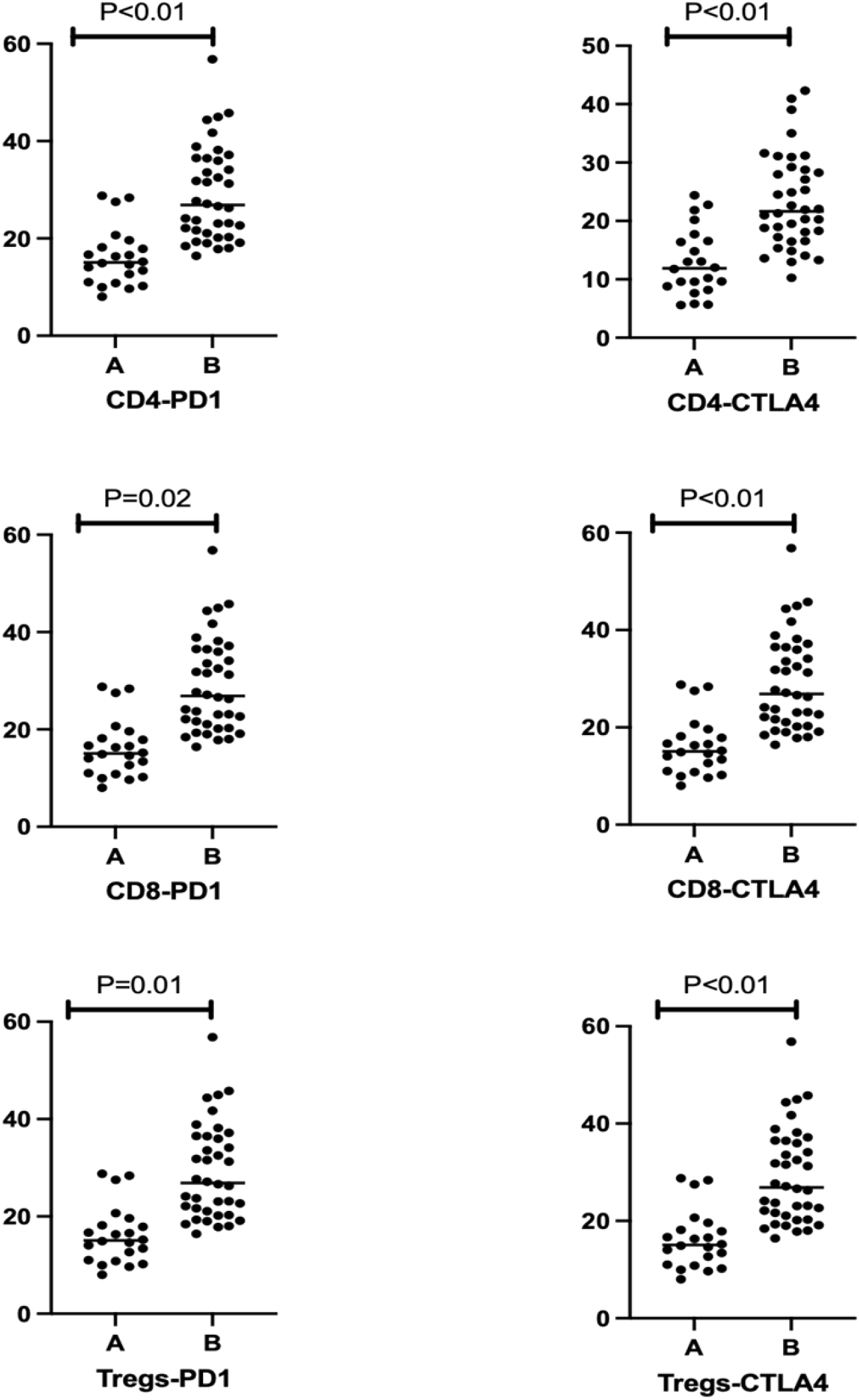
Level of PD-1/CTLA-4 in different nutritional statue

CD4^+^ PD-1, CD4^+^ CTLA-4, CD8^+^ PD-1, CD8^+^ CTLA-4, Tregs-PD-1 and Tregs-CTLA-4 were subjected to correlation analysis with the NRS-2002 score, BMI and ALB level, and the P values were statistically significant (Table 2).

## 4. Discussion

Many studies have reported the importance of nutritional status screening due to the associations of nutritional status with cardiovascular disease, cancer, mortality and other cause-related mortality risks^1-3,19,20^. Yanagita Y^21^ used the Geriatric Nutritional Risk Index (GNRI) as a tool to distinguish the interaction between nutritional status and aspiration pneumonia risk and concluded that the GNRI was higher in the survivor group than in the nonsurvivor group (78.1 vs. 68.6, P<0.01), as were the total protein (6.7 g/dL vs. 6.2 g/dL, P<0.01) and serum albumin levels (3.1 g/dL vs. 2.5 g/dL, P<0.01). He also calculated that the GNRI was an independent early predictor of mortality (OR = 0.383; 95% CI, 0.178–0.824; P < 0.05). However, we obtained similar results when we used the NRS-2002 score as a tool to assess nutritional status in our study. The number of hospital-free days (t=450, P<0.01), mortality and the occurrence of SCAP (X^2^=10.6, P<0.01) were lower in the no-risk group than in the malnutrition risk group. Moreover, we also found that the nutritional statue which indicated by NRS-2002 score was a risk factor for SCAP development. Both Iddir M and Yanagita Y emphasized the influence of ALB on the prognosis of pneumonia, but in the present study, ALB and BMI were not different between the two groups. However, many studies have shown that the ALB concentration is related to infectious disease risk. In combination with our previous study^22^, our findings indicate that nutritional risk screening and the level of ALB were correlated with the prognosis of severe pneumonia in elderly patients. The study also revealed that shock, invasive mechanical ventilation (IMV) or renal replacement therapy and the costs of the malnutrition risk group increased more severely than those of the no-risk group did. Therefore, we conclude that the NRS-2002 influences the prognosis of CAP patients.

T cells, including CD4^+^ T cells, CD8^+^ T cells and Tregs, are foundational to the immune system. PD-1 is often expressed in activated T lymphocytes and regulates T-cell activation and immune tolerance through binding with antigen-presenting cell (APC) ligands, resulting in T lymphocyte dysfunction, apoptosis and cytokine damage. CTLA-4 is an immunosuppressive factor that is expressed on the surface of activated T lymphocytes and generates signals that prevent T lymphocyte activation through competitively inhibiting ligand binding to CD28 and APCs. CD4^+^ and CD8^+^ T cells and Tregs, which are mediated by PD-1 and CTLA-4, are decreased in sepsis, while PD-1 and CTLA-4 are upregulated.^9-11^ Gong investigated the expression of PD-1 and CTLA-4 in CAP patients and reported that the percentages of PD-1 and CTLA-4 on Tregs were significantly greater than those in healthy controls. However, for both CD4^+^ and CD8^+^ T cells group, there were no differences between healthy individuals and CAP patients^12^. Unlike Gong’s research, PD-1 and CTLA-4 were related to the occurrence of SCAP and mortality in CD4^+^, CD8^+^ and Tregs in our study. However, when we performed a correlation test between PD-1/CTLA-4 and the PSI, we found that the results were positive, with higher PD-1 and CTLA-4 percentages indicating higher PSI grades. Therefore, we concluded that the percentage of PD-1/CTLA-4 levels were associated with the severity of CAP. Higher PD-1/CTLA-4 levels indicate more severe CAP. Malnutrition interferes with the suppression of immune reactions, but few studies have demonstrated this process at the molecular level. We conducted this process in patients with CAP and found that PD-1 and CTLA-4 expression was greater in the malnutrition risk group than in the no-risk group. Furthermore, we also demonstrated that the percentages of PD-1 and CTLA-4 were positively related to the NRS-2002 score. ALB and BMI are key indicators for evaluating nutritional status and are used to evaluate malnutrition^1^. In our research, the level of ALB was also related to PD-1 and CTLA-4 levels, whereas BMI was not correlated with these parameters.

Patients who had higher NRS-2002 scores or lower ALB levels presented higher PD-1 and CTLA-4 levels. Therefore, we propose that individuals who presented with malnutrition or hypoalbuminemia status may also have a high risk of immunosuppression in CAP patients. Tang W^23^ reported that the NRS-2002 score can predict the efficacy and prognosis of immunotherapy in patients with solid tumors treated with immune checkpoint inhibitor therapy. According to our research, the NRS-2002 score could also be an indicator of immunosuppression in patients with CAP and may be a prognostic factor in the treatment of CAP or other diseases.

## 5. Conclusion

A high NRS-2002 score may increase the degree of hospital stay and the occurrence of SCAP. Higher PD-1 and CTLA-4 levels were associated with a higher PSI grade. Nutritional status influenced the occurrence of immunosuppression, which is associated with the prognosis of CAP. Malnutrition status may increase the risk of immunosuppression, which is regulated by PD-1 and CTLA-4.

## Data Availability

All relevant data are within the manuscript and its Supporting Information files.

## Authors’contributions

Chenguang Zhang was responsible for data collection, literature reviewing and paper writing. Mingqiang Zhang and Xiangdong Mu conducted the experient and test the level of PD-1 and CTLA-4 in different T cell subgroups. Sheng Wu was responsible for paper writing and reviewing.

## Declarations

### Ethics approval and consent to participate

According the International Ethical Guidelines for Biomedical Research Involving Human Subjects (2002) and Helsinki Declaration (2013), Ethics Committee of the Beijing Tsinghua Changgung Hospital approved this research and all patients were informed the consent and agreed to take part in the research.

### Data availability statement

The datasets generated or analysed during the current study are not publicly available due the data will be taken for further research but are available from the corresponding author on reasonable request.

## Reference

1. Fernández-Lázaro D, Seco-Calvo J. Nutrition, Nutritional Status and Functionality. Nutrients. 2023 Apr 18;15(8):1944. doi: 10.3390/nu15081944. PMID: 37111162; PMCID: PMC10142726.

2. Mohajeri MH. Nutrition and Aging. Int J Mol Sci. 2023 May 25;24(11):9265. doi: 10.3390/ijms24119265. PMID: 37298216; PMCID: PMC10253359.

3. Frederiks P, Peetermans M, Wilmer A. Nutritional support in the cardiac intensive care unit. Eur Heart J Acute Cardiovasc Care. 2024 May 7;13(4):373–379. doi: 10.1093/ehjacc/zuae018. PMID: 38333990.

4. Chen LK, Arai H, Assantachai P, Akishita M, Chew STH, Dumlao LC, Duque G, Woo J. Roles of nutrition in muscle health of community-dwelling older adults: evidence-based expert consensus from Asian Working Group for Sarcopenia. J Cachexia Sarcopenia Muscle. 2022 Jun;13(3):1653–1672. doi: 10.1002/jcsm.12981. Epub 2022 Mar 20. PMID: 35307982; PMCID: PMC9178363.

5. Karagiannidis C, Krause F, Bentlage C, Wolff J, Bein T, Windisch W, Busse R. In-hospital mortality, comorbidities, and costs of one million mechanically ventilated patients in Germany: a nationwide observational study before, during, and after the COVID-19 pandemic. Lancet Reg Health Eur. 2024 Jun 7;42:100954. doi: 10.1016/j.lanepe.2024.100954. PMID: 39070745; PMCID: PMC11281923.

6. Iddir M, Brito A, Dingeo G, Fernandez Del Campo SS, Samouda H, La Frano MR, Bohn T. Strengthening the Immune System and Reducing Inflammation and Oxidative Stress through Diet and Nutrition: Considerations during the COVID-19 Crisis. Nutrients. 2020 May 27;12(6):1562. doi: 10.3390/nu12061562. PMID: 32471251; PMCID: PMC7352291.

7. Meydani SN, Beck MA. The relationship of host nutritional status to immune function. Mol Aspects Med. 2012 Feb;33(1):1. doi: 10.1016/j.mam.2011.12.001. Epub 2011 Dec 14. PMID: 22196799.

8. Sümer A, Uzun LN, Özbek YD, Tok HH, Altınsoy C. Nutrition improves COVID-19 clinical progress. Ir J Med Sci. 2022 Oct;191(5):1967–1972. doi: 10.1007/s11845-021-02868-w. Epub 2022 Jan 15. PMID: 35031937; PMCID: PMC8760115.

9. Wu L, Liu H, Liu H. Prognostic significance of PD-1, CTLA-4, CD4, and CD8 expression in olfactory neuroblastoma. Clin Neuropathol. 2023 Mar-Apr;42(2):47–53. doi: 10.5414/NP301519. PMID: 36708210.

10. Rowshanravan B, Halliday N, Sansom DM. CTLA-4: a moving target in immunotherapy. Blood. 2018 Jan 4;131(1):58–67. doi: 10.1182/blood-2017-06-741033. Epub 2017 Nov 8. PMID: 29118008; PMCID: PMC6317697.

11. Gao DN, Yang ZX, Qi QH. Roles of PD-1, Tim-3 and CTLA-4 in immunoregulation in regulatory T cells among patients with sepsis. Int J Clin Exp Med. 2015 Oct 15;8(10):18998-9005. PMID: 26770525; PMCID: PMC4694425.

12. Gong H, Zhao J, Xu W, Wan Y, Mu X, Zhang M. The distribution of myeloid-derived suppressor cells subsets and up-regulation of programmed death-1/PD-L1 axis in peripheral blood of adult CAP patients. PLoS One. 2023 Sep 27;18(9):e0291455. doi: 10.1371/journal.pone.0291455. PMID: 37756307; PMCID: PMC10529571.

13. Pletz MW, Blasi F, Chalmers JD, Dela Cruz CS, Feldman C, Luna CM, Ramirez JA, Shindo Y, Stolz D, Torres A, Webb B, Welte T, Wunderink R, Aliberti S. International Perspective on the New 2019 American Thoracic Society/Infectious Diseases Society of America Community-Acquired Pneumonia Guideline: A Critical Appraisal by a Global Expert Panel. Chest. 2020 Nov;158(5):1912–1918. doi: 10.1016/j.chest.2020.07.089. Epub 2020 Aug 25. PMID: 32858009; PMCID: PMC7445464.

14. Nair GB, Niederman MS. Updates on community acquired pneumonia management in the ICU. Pharmacol Ther. 2021 Jan;217:107663. doi: 10.1016/j.pharmthera.2020.107663. Epub 2020 Aug 15. PMID: 32805298; PMCID: PMC7428725.

15. Metlay JP, Waterer GW, Long AC, Anzueto A, Brozek J, Crothers K, Cooley LA, Dean NC, Fine MJ, Flanders SA, Griffin MR, Metersky ML, Musher DM, Restrepo MI, Whitney CG. Diagnosis and Treatment of Adults with Community-acquired Pneumonia. An Official Clinical Practice Guideline of the American Thoracic Society and Infectious Diseases Society of America. Am J Respir Crit Care Med. 2019 Oct 1;200(7):e45–e67. doi: 10.1164/rccm.201908-1581ST. PMID: 31573350; PMCID: PMC6812437.

16. Mandell LA. Community-acquired pneumonia: An overview. Postgrad Med. 2015 Aug;127(6):607–15. doi: 10.1080/00325481.2015.1074030. PMID: 26224210; PMCID: PMC7103686.

17. Zhang Z, Wan Z, Zhu Y, Zhang L, Zhang L, Wan H. Prevalence of malnutrition comparing NRS2002, MUST, and PG-SGA with the GLIM criteria in adults with cancer: A multi-center study. Nutrition. 2021 Mar;83:111072. doi: 10.1016/j.nut.2020.111072. Epub 2020 Nov 19. PMID: 33360034.

18. Kondrup J, Allison SP, Elia M, Vellas B, Plauth M; Educational and Clinical Practice Committee, European Society of Parenteral and Enteral Nutrition (ESPEN). ESPEN guidelines for nutrition screening 2002. Clin Nutr. 2003 Aug;22(4):415–21. doi: 10.1016/s0261-5614(03)00098-0. PMID: 12880610.

19. Mirizzi, A.; Aballay, L.R.; Misciagna, G.; Caruso, M.G.; Bonfiglio, C.; Sorino, P.; Bianco, A.; Campanella, A.; Franco, I.; Curci, R.; et al. Modified WCRF/AICR Score and All-Cause, Digestive System, Cardiovascular, Cancer and Other-Cause-Related Mortality: A Competing Risk Analysis of Two Cohort Studies Conducted in Southern Italy. Nutrients 2021, 13, 4002.

20. Mugica-Errazquin, I.; Zarrazquin, I.; Seco-Calvo, J.; Gil-Goikouria, J.; Rodriguez-Larrad, A.; Virgala, J.; Arizaga, N.; Matilla-Alejos, B.; Irazusta, J.; Kortajarena, M. The Nutritional Status of Long-Term Institutionalized Older Adults Is Associated with Functional Status, Physical Performance and Activity, and Frailty. Nutrients 2021, 13, 3716.

21. Yanagita Y, Arizono S, Tawara Y, Oomagari M, Machiguchi H, Yokomura K, Katagiri N, Iida Y. The severity of nutrition and pneumonia predicts survival in patients with aspiration pneumonia: A retrospective observational study. Clin Respir J. 2022 Jul;16(7):522–532. doi: 10.1111/crj.13521. Epub 2022 Jul 5. PMID: 35789107; PMCID: PMC9329015.

22. Zhang CG, Chen XY, Zhang XY, et al. Correlation between nutritional risk screening and the prognosis of elderly severe pneumonia. Chin J Crit Care Med, 2023, 43 (03): 175–179.

23. Tang W, Li C, Huang D, Zhou S, Zheng H, Wang Q, Zhang X, Fu J. NRS2002 score as a prognostic factor in solid tumors treated with immune checkpoint inhibitor therapy: a real-world evidence analysis. Cancer Biol Ther. 2024 Dec 31;25(1):2358551. doi: 10.1080/15384047.2024.2358551. Epub 2024 May 30. PMID: 38813753; PMCID: PMC11141475.

